# Magnitude of Perinatal Mortality in Sub-Saharan Africa: A Systematic Review and Meta-Analysis of Observational Studies

**DOI:** 10.1101/2025.09.16.25335956

**Authors:** Musonda Makasa, Patrick Kaonga, Choolwe Jacobs, Kabilwa Silumesi, Mercy Wamunyima Monde, Bellington Vwalika

## Abstract

**Background:** In 2023, an estimated 2.3 million newborns died globally, nearly half of all under-five deaths. Although global perinatal mortality has declined, sub-Saharan Africa continues to carry a disproportionate burden. This study aimed to determine the magnitude and synthesize pooled estimates of predictors of perinatal mortality in sub-Saharan Africa.

**Methods:** We systematically searched PubMed, EMBASE, Scopus, CINAHL, ProQuest Global, IRIS, and Google Scholar for observational studies published in English between January 2000 and December 2024. Study quality and risk of bias were appraised independently using the JBI and STROBE checklists. Data were analysed in Stata 17. Heterogeneity was assessed with I² and Q-statistics, and random-effects models generated pooled estimates with 95% confidence intervals (CI). Publication bias could not be formally assessed due to the small number of studies per predictor.

**Results:** Fourteen studies met inclusion criteria. Most were of moderate to high quality, with confounding and incomplete follow-up being the most frequent risks of bias. Pooled odds ratios (ORs) showed significant associations between perinatal mortality and parity (3.96; 95% CI: 2.10– 7.47), rural residence (2.56; 95% CI: 1.68–3.91), referral status (6.90; 95% CI: 2.71–17.52), antepartum haemorrhage (7.28; 95% CI: 2.70–19.65), congenital anomalies (9.70; 95% CI: 3.50– 27.00), induced labour (2.19; 95% CI: 1.46–3.29), dystocia (3.86; 95% CI: 2.26–6.60), non-use of the partograph (3.86; 95% CI: 2.26–6.60), prematurity (2.77; 95% CI: 1.81–4.26), and low birthweight (3.04; 95% CI: 0.98–9.46). A history of perinatal mortality was also predictive (RR 3.37; 95% CI: 1.36–8.34). One unpooled study reported syphilis, Rhesus status, and foetal presentation as additional risks.

**Conclusion:** Sociodemographic factors, maternal and obstetric complications, and infections emerged as key predictors of perinatal mortality in sub-Saharan Africa. While most included studies were of moderate to high quality, residual bias related to confounding and follow-up was noted. These findings underscore the multifactorial nature of perinatal mortality and highlight the need for integrated maternal and newborn health interventions to accelerate progress toward global targets.

**Systematic review registration: PROSPERO CRD42023437432**

## Introduction

Perinatal mortality is a critical global health challenge, particularly in low-income regions, where it imposes substantial economic, social, and emotional burdens on women, families, and society [1, 2]. In 2023, an estimated 2.3 million newborns died globally, accounting for nearly half of all under-five deaths—equivalent to approximately 6,300 neonatal deaths each day [3, 4]. The majority of these deaths occurred in low- and middle-income countries (LMICs), particularly in sub-Saharan Africa (SSA) and South Asia, where access to quality maternal and newborn healthcare remains limited [3]. Perinatal mortality, which encompasses stillbirths, is defined as foetal deaths after 28 weeks of gestation, and early neonatal deaths occurring within the first seven days of life. When gestational age is uncertain, births weighing 1,000 grams or more are included, in line with WHO recommendations for international comparisons [5].

The launch of the Millennium Development Goals (MDGs) and, later, the Sustainable Development Goals (SDGs) contributed to gradual global reductions in perinatal mortality [6]. SDG target 3.2 specifically aims to reduce neonatal mortality to at least 12 per 1,000 live births by 2030 [7]. Between 2000 and 2015, global perinatal mortality declined from 5.7 million to 4.1 million deaths, with 95% of these occurring in South Asia and SSA, while stillbirth rates dropped from 24.7 to 18.4 per 1,000 live births [8, 9]. Despite these gains, disparities persist: high-income countries account for fewer than 2% of stillbirths, while approximately 98% occur in LMICs, with SSA and South Asia contributing nearly 77% of cases [10, 11]. Despite a 30–33% decline in stillbirth and neonatal mortality rates, SSA still accounts for 47% of global stillbirths and 46% of newborn deaths, and is unlikely to meet the SDG neonatal mortality target by 2030 at the current rate of progress [12, 13]. Akombi and Renzaho (9) reported that SSA had the highest perinatal mortality rate globally (42.95 per 1,000 live births), with Nigeria and Ethiopia among the worst affected.

Perinatal mortality is widely regarded as an indicator of the quality of antenatal, intrapartum, and early neonatal care in a given setting [14]. The causes of perinatal mortality are multifactorial, encompassing interactions among maternal lifestyle, medical and obstetric complications, and broader environmental and health systems factors. A recent analysis highlights that maternal and perinatal outcomes depend on this intricate interplay, shaped by both biomedical and contextual health system factors [15]. Barriers to reducing perinatal mortality in SSA include inadequate access to quality and equitable care, shortages of skilled providers, and community- and geographic-level barriers to timely facility-based services. Efforts to address these challenges include Every Newborn Action Plan (ENAP), perinatal audit initiatives, and other global strategies such as the UN Secretary-General’s Every Woman Every Child framework, UNICEF’s Every Child Alive campaign, and the WHO-led Quality of Care Network [2, 16, 17]. Although global initiatives such as the MDGs and SDGs have driven some reductions in perinatal and neonatal mortality, progress in SSA remains slow and uneven.

Understanding the predictors of perinatal mortality in sub-Saharan Africa is therefore critical for informing targeted interventions and policies to address this persistently high burden. This study undertook a systematic review and meta-analysis of observational studies published between 2000 and 2023, quantifying pooled prevalence, odds ratios, and relative risks of perinatal mortality across the region. By synthesizing evidence from diverse study designs and country contexts, it aimed to generate robust and specific insights into the social, maternal, obstetric, and neonatal predictors of perinatal mortality. These findings are intended to strengthen the evidence base for context-appropriate strategies, health system improvements, and accelerated progress toward global maternal and newborn health targets in SSA.

## Methods and Materials

This systematic review and meta-analysis were conducted and reported in accordance with the PRISMA 2020 guidelines [18]. The protocol was registered prospectively with PROSPERO (CRD42023437432).

## Inclusion and exclusion criteria

### Inclusion Criteria

Eligible studies were observational in design (cohort, case–control, and cross-sectional) and published between January 2000 and December 2023. Studies had to be conducted in sub-Saharan Africa and report on stillbirth, early neonatal mortality (ENND), or perinatal mortality, with sufficient data to extract or compute effect estimates.

We excluded audits, systematic reviews, qualitative reports, abstracts, case reports, lacked sample justification, conference presentations, and expert opinions. Studies conducted outside SSA, published outside the prespecified date range, or failing to provide sufficient outcome data were also excluded.

### Search strategy

An electronic systematic search was conducted in PubMed (n = 2,741), with the strategy adapted for EMBASE (n = 75), Scopus (n = 360), and CINAHL (n = 174). Grey literature sources were also explored, including ProQuest Global Theses and Dissertations (n = 23), WHO IRIS (n = 1,017), and Google Scholar (first 200 results screened). In addition, reference lists of included articles were hand-searched to identify any studies that may have been missed. The final search was completed on December 21, 2023.

To ensure a balance between precision and sensitivity, the strategy incorporated Medical Subject Headings (MeSH) and free-text terms combined with Boolean operators (“AND,” “OR”), truncation, and synonyms. Core search terms included *perinatal mortality, perinatal death, stillbirth, intrauterine fetal/foetal death, early neonatal death, early neonatal mortality, incidence, prevalence, ratio, risk factors, rate, magnitude,* and *sub-Saharan Africa* (or *Africa south of the Sahara*, *SSA*). The search strategy was iteratively developed and jointly reviewed by the research team to ensure completeness and reproducibility. The search strategy details for perinatal mortality and the associated risk factors is illustrated in the supplementary (see S1 table for ease of reference).

### Study identification and selection

All retrieved citations were imported into EndNote referencing software and then screened in Rayyan.ai. Four reviewers (MM, BN, KC, JS) independently screened titles and abstracts, followed by full-text screening by three reviewers (MM, KS, MK). Disagreements were resolved by consensus. Reasons for exclusion at the full-text stage were recorded, and the overall selection process is illustrated in a PRISMA flow diagram (Figure 1).

**Figure 1:**
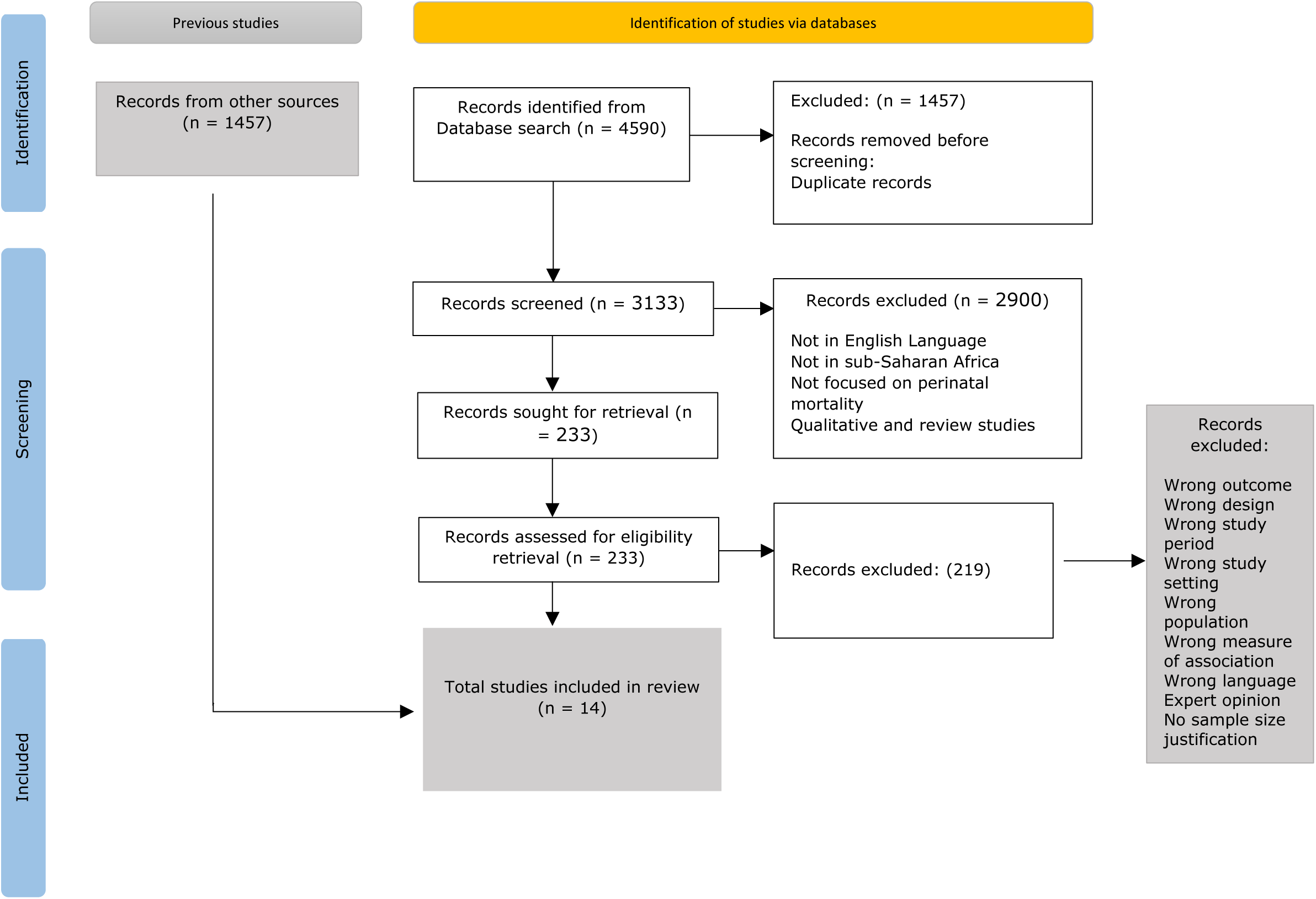
The figure illustrates the flow diagram of search and study selection process, according to PRISMA, for perinatal mortality in sub-Saharan Africa from January 2000 to December 2023.

### Risk of Bias assessment

The methodological quality of included studies was assessed independently by two reviewers (BN, KC) using the Joanna Briggs Institute (JBI) critical appraisal checklists for cohort, case–control, and cross-sectional studies [19, 20]. The JBI Critical Appraisal Checklist is provided in Supplementary Table 2 (S2 Table). Although the STROBE checklist was consulted for reporting quality, it was not used as a formal risk of bias tool. Results of the quality appraisal informed sensitivity analyses.

### Data abstraction

Data extraction was performed using a standardized JBI-based form. Extracted variables included study characteristics (author, year, country, design, setting, population), exposure and outcome definitions, sample sizes, and reported effect estimates (Odds Ratio (ORs), Risk Ratio (RRs), prevalence rates)). Discrepancies were resolved through discussion.

### Data synthesis and Statistical Analysis

Data from eligible studies were cleaned and formatted in Microsoft Excel, then exported into Stata version 17 for meta-analysis. Study-level prevalence estimates were transformed using the Freeman–Tukey double arcsine method to stabilize variance. ORs and RRs were log-transformed prior to pooling. Between-study heterogeneity was first assessed visually with forest plots and quantified using the I² statistic, with thresholds of 25%, 50%, and 75% representing low, moderate, and high heterogeneity, respectively. Pooled effect sizes—including prevalence, ORs, and RRs— were calculated using a random-effects model (DerSimonian–Laird method), with τ² estimating between-study variance. Sensitivity analyses were performed to identify studies that disproportionately contributed to heterogeneity.

### Subgroup and Sensitivity Analysis

Subgroup analyses were prespecified by study design (cohort, case–control, cross-sectional) and geographic region, with additional exploratory analyses conducted by year of publication and study setting. Owing to the small number of studies in several categories, subgroup findings were interpreted with caution.

A total of 14 studies were included in the quantitative synthesis. Because study designs and reported predictors varied, common predictors were identified within each design and pooled accordingly. Among cross-sectional studies (n = 2), parity was the only consistently reported predictor and was analysed as a determinant of perinatal mortality. For cohort studies (n = 5), two studies each contributed data on history of stillbirth, intrauterine foetal death (IUFD), and early neonatal death (ENND). The case–control studies (n = 7) formed the largest group; within these, two studies each were meta-analysed for rural residence, referral status, antepartum haemorrhage (APH), foetal congenital anomalies, induced labour, dystocia, non-use of the partograph, and prematurity. Low birthweight (LBW) emerged as a key predictor, with five studies contributing to the pooled analysis.

Sensitivity analyses indicated that the pooled estimates were robust. Excluding lower-quality studies did not substantially alter the effect sizes. Results remained consistent under the primary random-effects model, and adjustment for potential publication bias using the trim-and-fill method did not materially change the direction or magnitude of the associations.

### Risk of Bias Assessment

We planned to assess publication bias using funnel plots and asymmetry and Egger’s regression test. However, this was not performed, as each meta-analysis included fewer than the recommended minimum of 10 studies, consistent with Cochrane Handbook guidance [21]. Final outputs included pooled prevalence estimates and pooled effect measures (ORs and RRs), providing a quantitative summary of perinatal mortality and its predictors across sub-Saharan Africa.

### Certainty of Evidence

The certainty of the evidence was assessed using the GRADE (Grading of Recommendations Assessment, Development and Evaluation) approach. We evaluated each outcome across five domains: risk of bias, inconsistency, indirectness, imprecision, and publication bias. Based on these assessments, we assigned a level of confidence (high, moderate, low, or very low) to the pooled estimates of perinatal mortality and to the identified predictors. This framework allowed us to systematically judge the overall strength of the evidence and to highlight areas where limitations reduced confidence in the findings.

### Ethics Consideration

This review and included was based on analysis of previously published studies and publicly accessible data sources. No new data were collected directly from human participants, and therefore institutional ethical approval and informed consent were not required. The review protocol was prospectively registered in PROSPERO (CRD42023437432).

### Results

A total of 4590 articles were retrieved through electronic searches by titles and abstracts. Of these records, we arrived at a total of 14 eligible studies to estimate the observed and adjusted effect sizes, the extent of heterogeneity, and the existence of publication bias for perinatal mortality and the selected predictors.

## Description of Studies

A total of 14 studies were included in this review, comprising two cross-sectional studies, seven case-control studies, and five cohort studies. In terms of study setting, one was a household survey, one was conducted in a public health centre, two in public hospitals, two in urban hospitals, and eight in tertiary hospitals.

Geographically, the studies spanned different regions of sub-Saharan Africa. From West Africa, two studies were identified: one from Burkina Faso [22] and another from Ghana [23]. From Central Africa, all three originated from Cameroon [24–26]. In East Africa, one study was from Sudan [27], four from Ethiopia [28–31], three from Uganda [32–34], and one from Rwanda [35].

The review covered articles published between January 2000 and December 2023. Sample sizes ranged from 341 participants in the study by Musooko, Kakaire (32), which recorded the smallest sample, to 2,279 participants in the study by Agena and Modiba (29) in Ethiopia, which had the largest. In total, the studies included 11,845 pregnant women across the region. Outcomes varied: stillbirths were assessed in nine studies, perinatal death in four, and early neonatal death in one. A broad range of maternal, obstetric, and neonatal predictors were reported. Sociodemographic predictors included rural residence [28, 31, 35] and maternal age [34, 35].

Obstetric complications such as antepartum haemorrhage [30], referral status [24, 25], and prolonged or complicated labour [23, 24] were also frequently highlighted. Neonatal predictors, particularly low birthweight and prematurity, emerged consistently across several studies [23–26, 30, 31]. A detailed summary of study characteristics—including author, year, study design, sample size, outcomes assessed, and key predictors—is provided in Table 1 after the Figure 1 below, which presents the PRISMA flow diagram outlining the process of study identification, screening, eligibility assessment, and inclusion

**Table 1:**
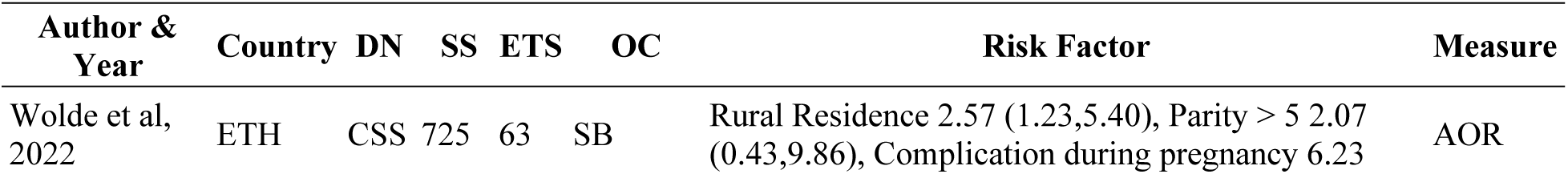
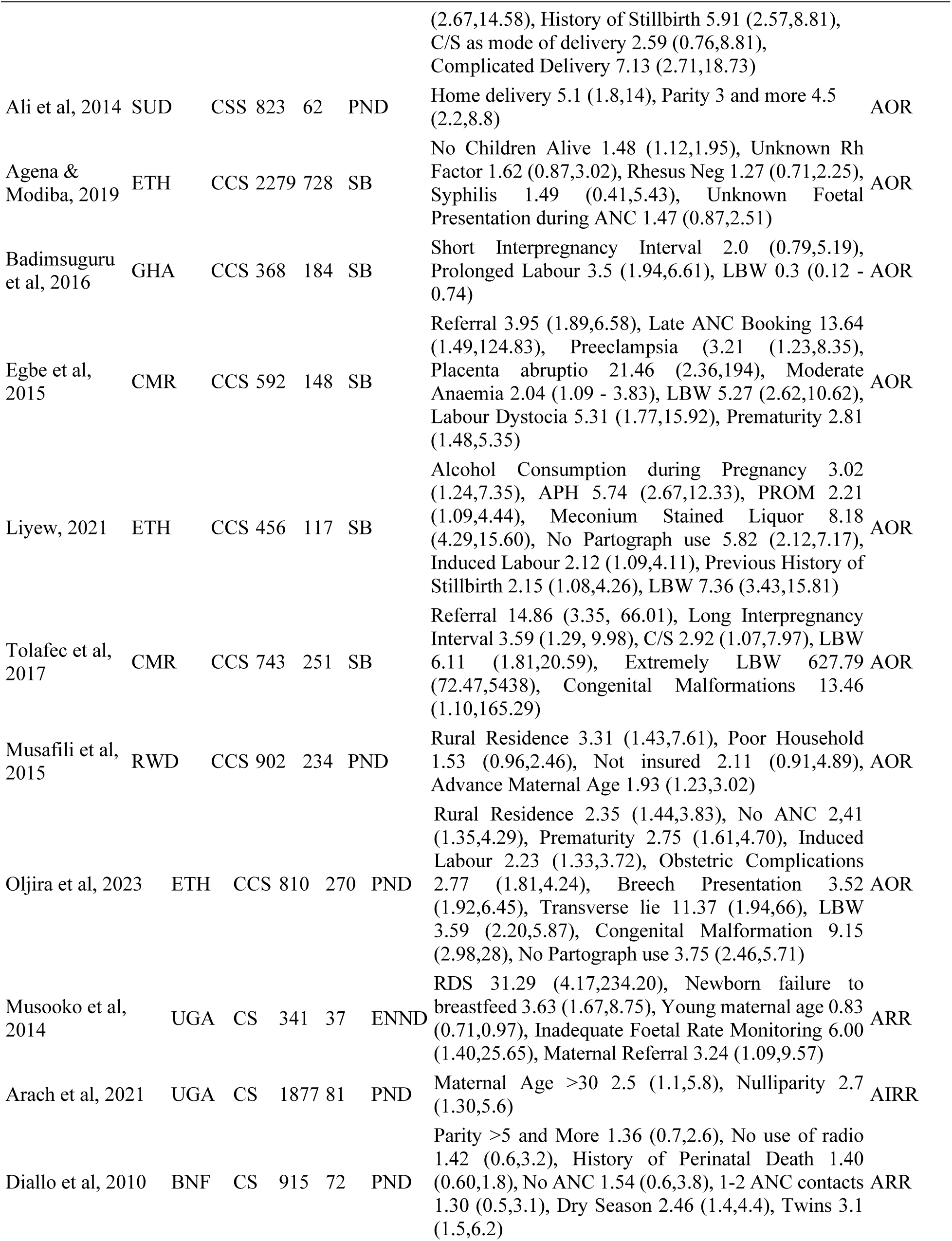
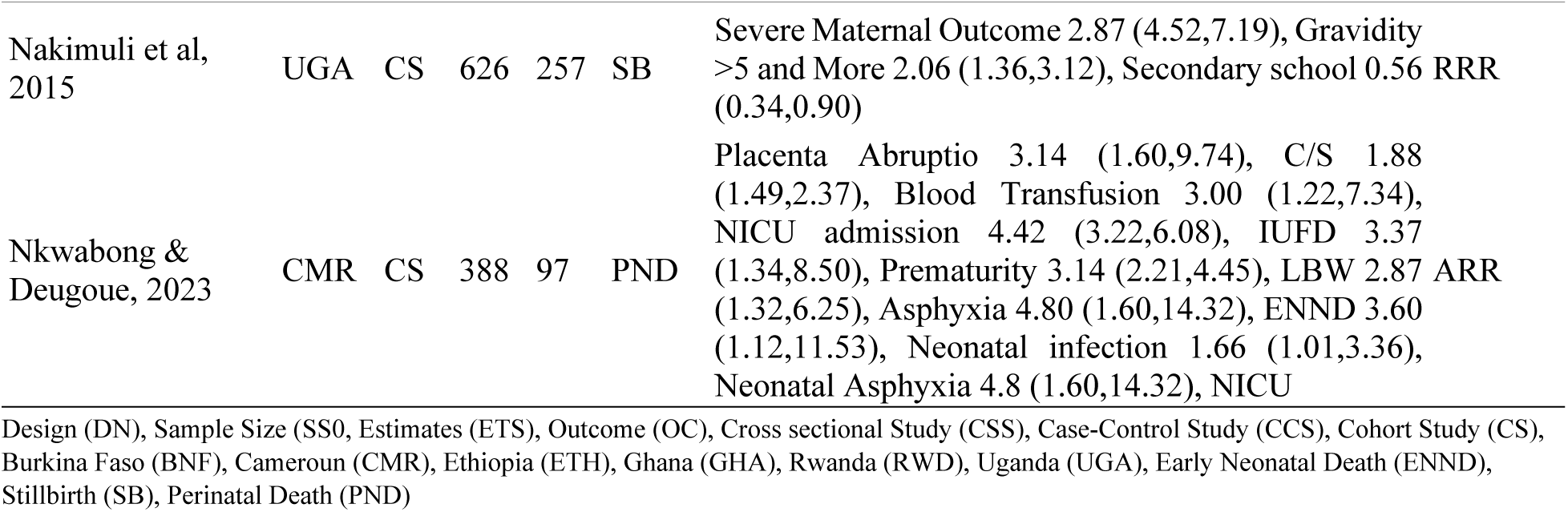
Baseline Characteristics of Studies included in the Systematic Review of Perinatal Mortality in SSA (Jan 2000 – Dec 2023)

### Selection of studies

The following is a flow chart of illustrating the number of articles screened and retrieved.

### Study Quality and Risk of Bias Assessment

When we assessed the quality of the studies included in this review using the Joanna Briggs Institute (JBI) critical appraisal tools, we found that most were of moderate to high quality. Six studies were rated as high quality with a low risk of bias, six were judged moderate, and two showed lower quality. Across nearly all studies, the measurement of exposures, outcomes, and statistical methods was robust. However, a common weakness was the handling of confounders— some studies did not clearly identify or adjust for them. In the cohort studies, another area of concern was follow-up, where the duration or completeness of tracking participants was not always clearly reported.

Cross-sectional studies, while valuable, tended to provide less detail on how they dealt with confounding, and this limited their quality rating. Case-control studies generally performed better, especially those from tertiary hospitals where exposures and outcomes were measured more rigorously. Cohort studies stood out for their stronger design and analyses, though some still left questions about attrition. Taken together, these findings show that the evidence base is reasonably strong but not without limitations. The main areas that could affect interpretation relate to confounding and incomplete follow-up. Despite this, the overall quality of the evidence was sufficient to support our pooled analyses. To ease reference, see the supplementary table S2, critical appraisal of included studies.

### Meta-analysis

A total of 14 studies were included in the quantitative synthesis. Since the studies varied in design and reported different predictors, common predictors were identified and meta-analysed within each study type.

- Cross-sectional studies (n = 2): Parity was the only consistently reported predictor and was pooled as a predictor of perinatal mortality.
- Cohort studies (n = 5): Two studies each were pooled for history of stillbirth, IUFD, and early neonatal death ENND.
- Case-control studies (n = 7): The largest group. Pooled analyses were conducted using two studies each for rural residence, referral status, APH, foetal congenital anomalies, induced labour, dystocia, non-use of the partograph, and prematurity.
- LBW: This was the most frequently reported predictor, with five studies contributing to the pooled analysis.

The pooled prevalence of perinatal mortality, as well as pooled ORs and RRs for predictors of perinatal mortality, are summarised and illustrated in Figures 2–12. Subgroup analyses by study design and geographic region were attempted to explore sources of heterogeneity; however, interpretation was constrained by the small number of studies in some subgroups.

**Fig 2:**
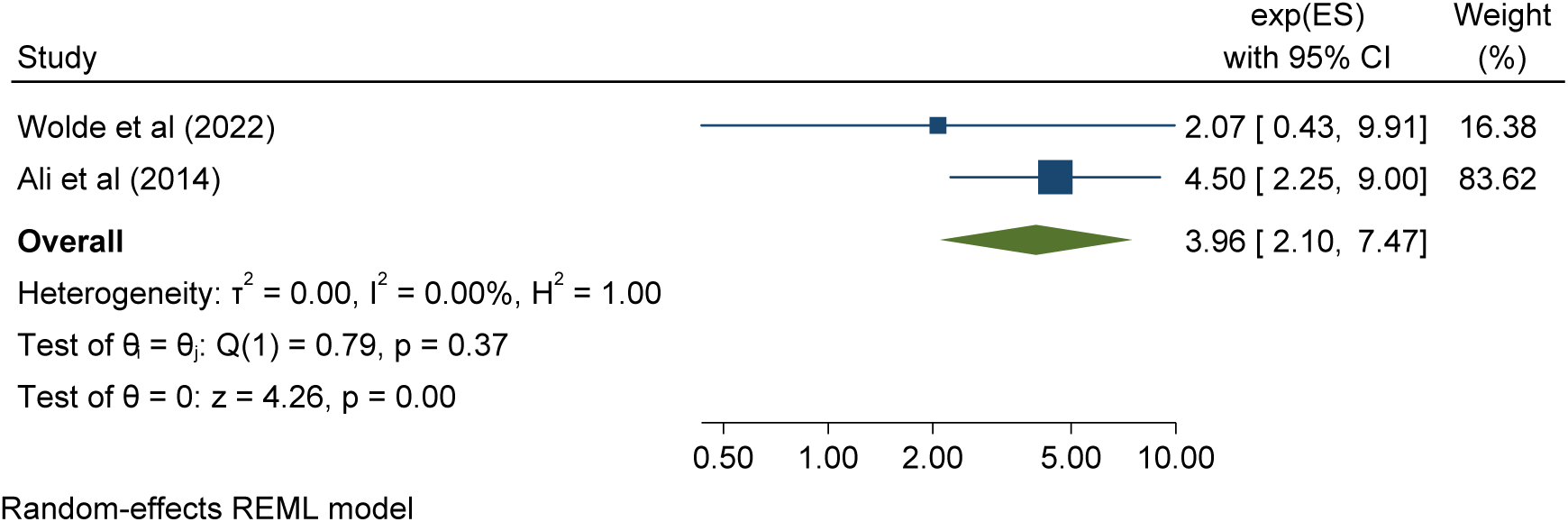
Pooled prevalence for parity.

**Fig 3:**
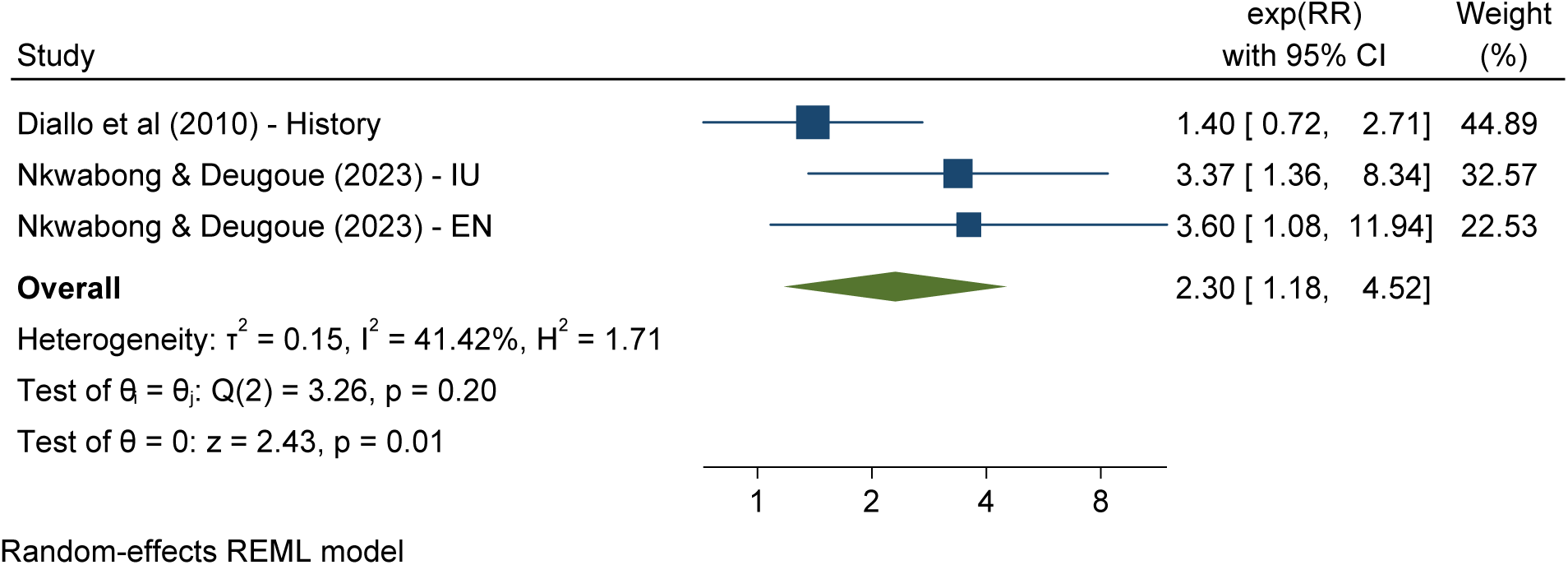
Pooled risk ratio for perinatal mortality.

**Fig 4:**
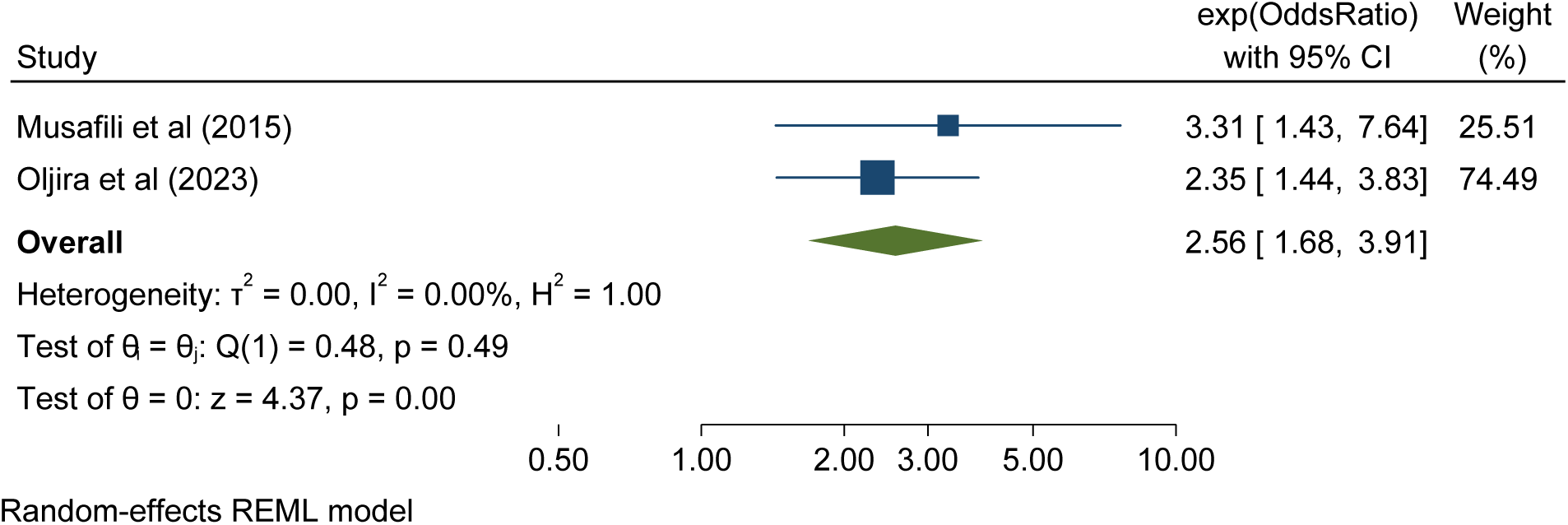
Pooled OR for rural residence.

**Fig 5:**
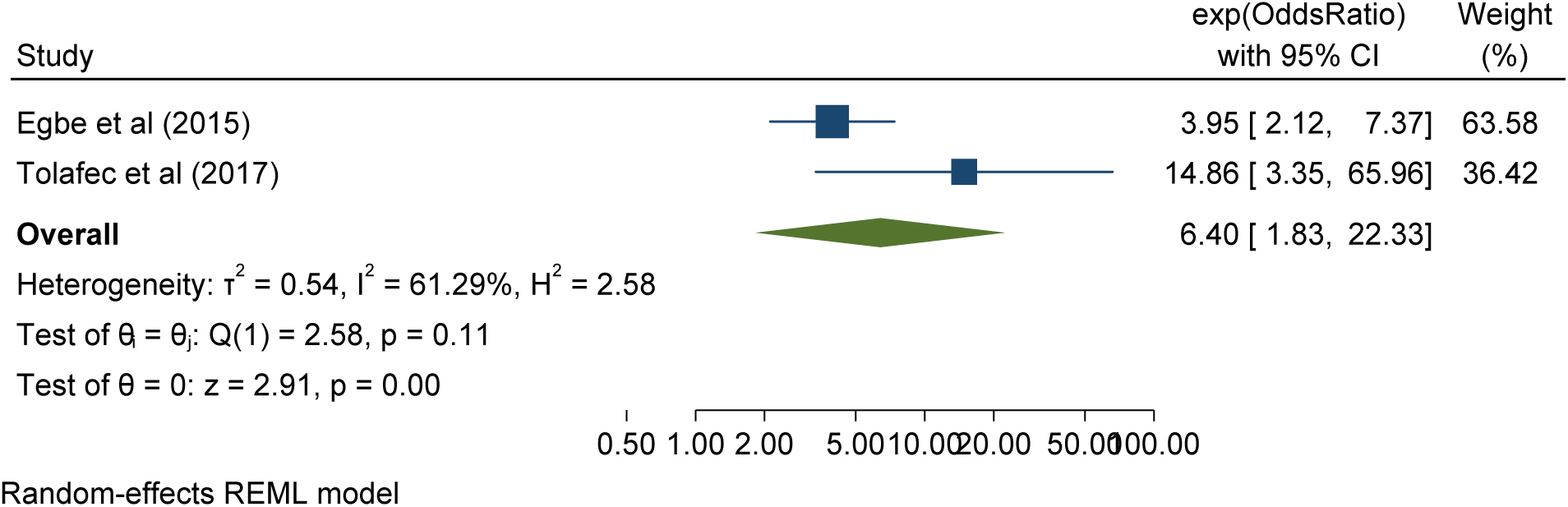
Pooled OR for referral status.

**Fig 6:**
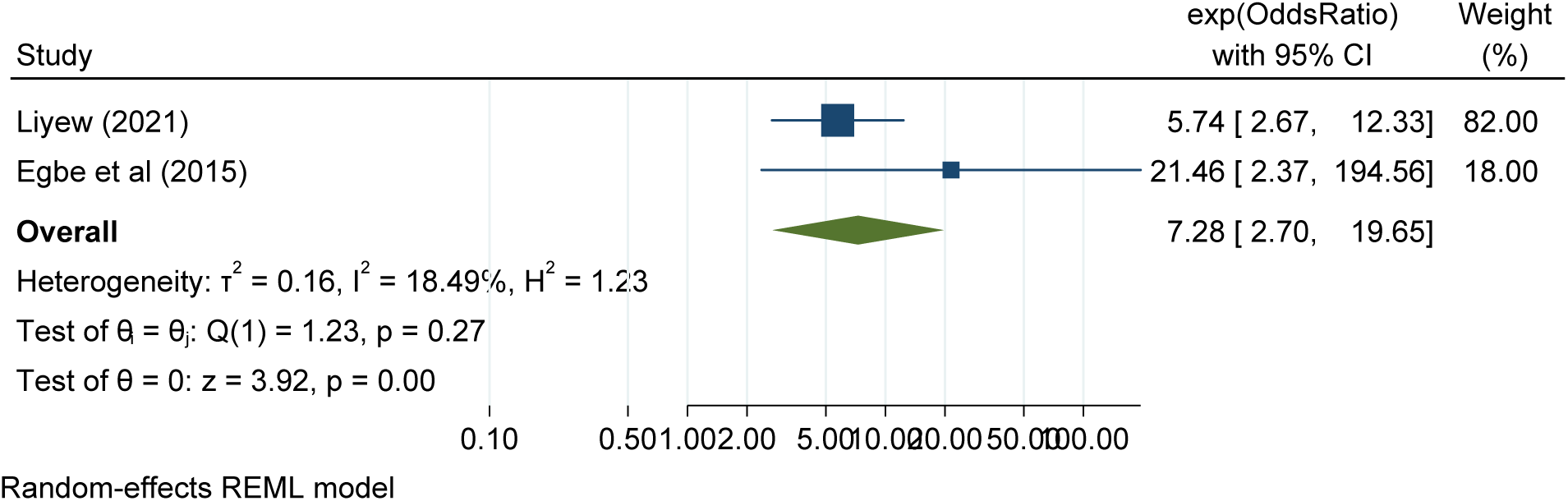
Pooled OR for APH.

**Fig 7:**
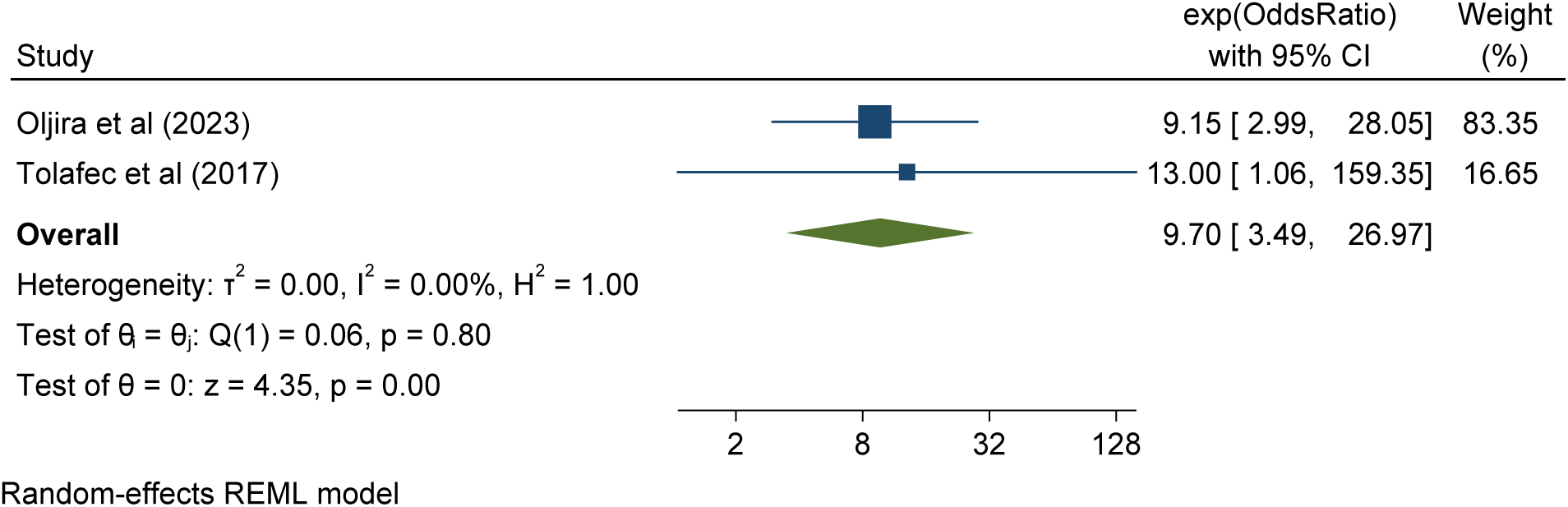
Pooled OR for congenital anomalies.

**Fig 8:**
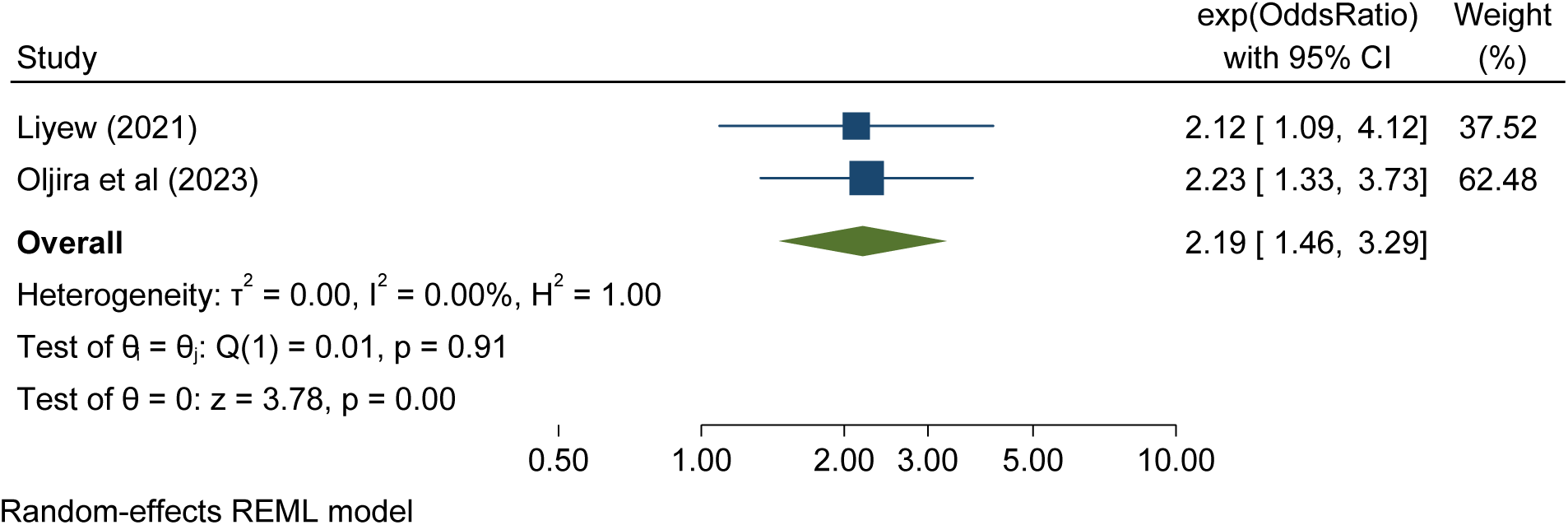
Pooled OR for induced labour.

**Fig 9:**
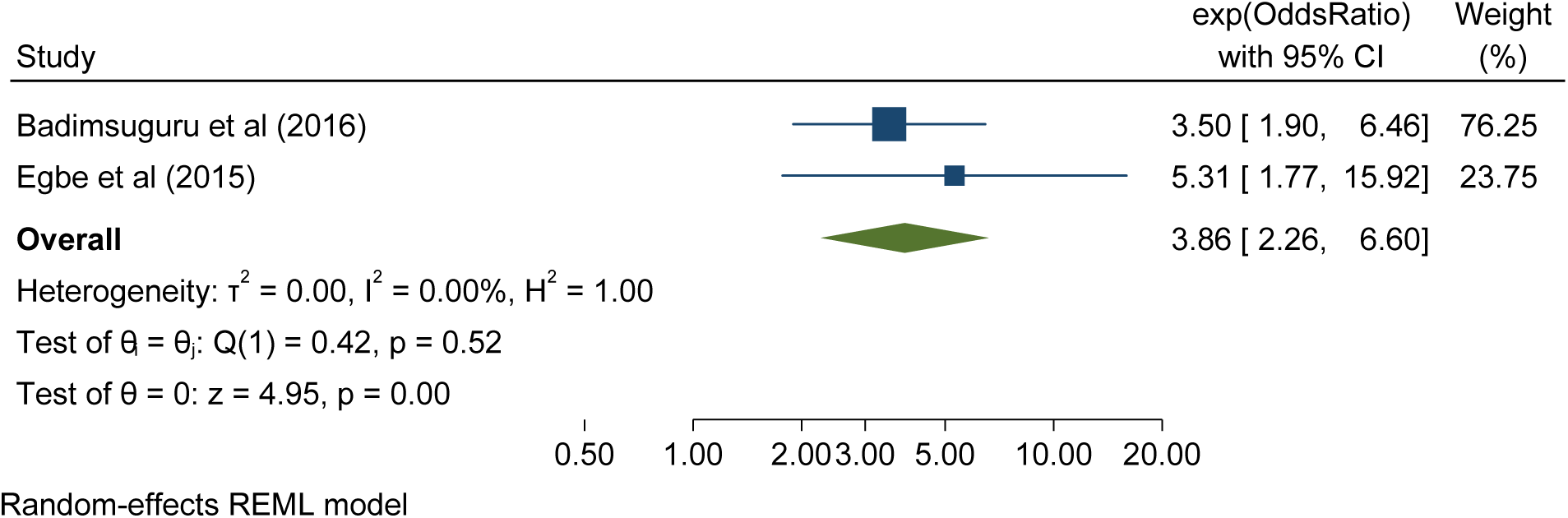
Pooled OR for dystocia.

**Fig 10.**
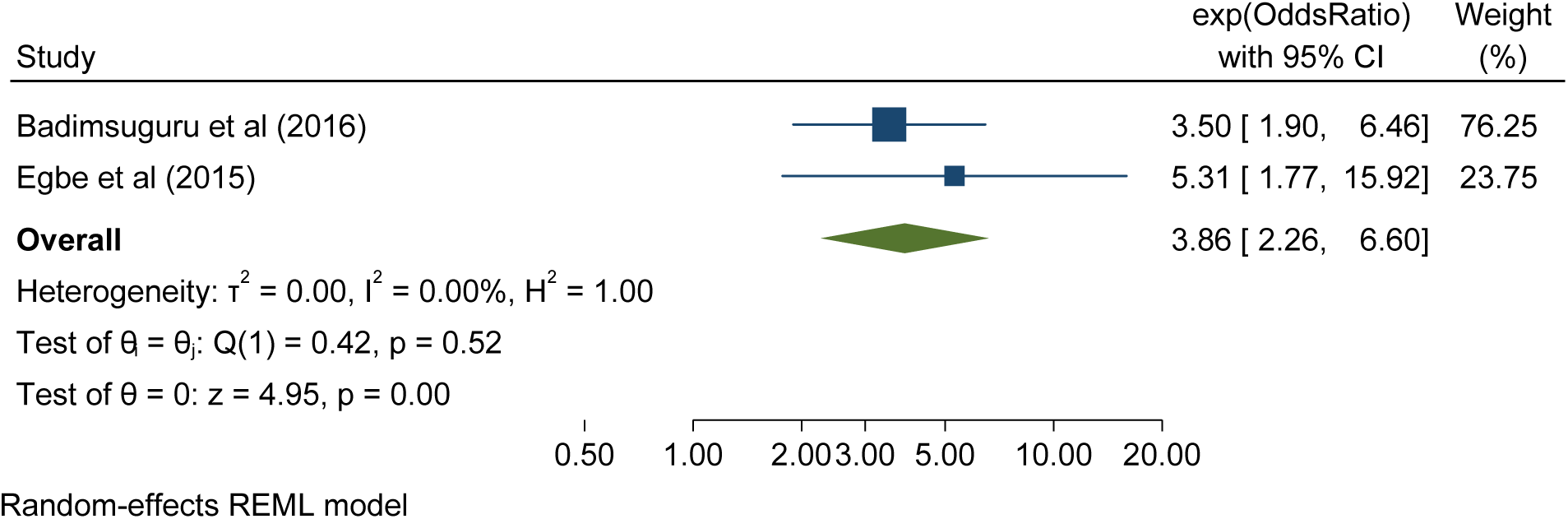
Pooled for non-use of the partograph.

**Fig 11.**
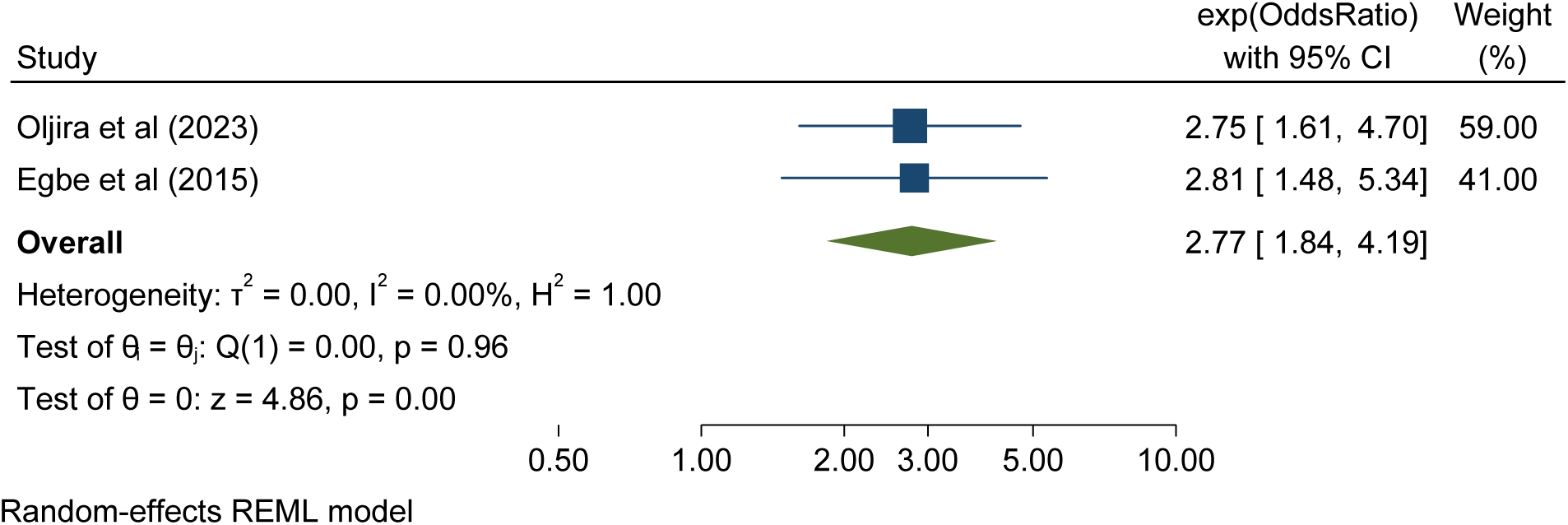
Pooled OR for prematurity.

**Fig 12:**
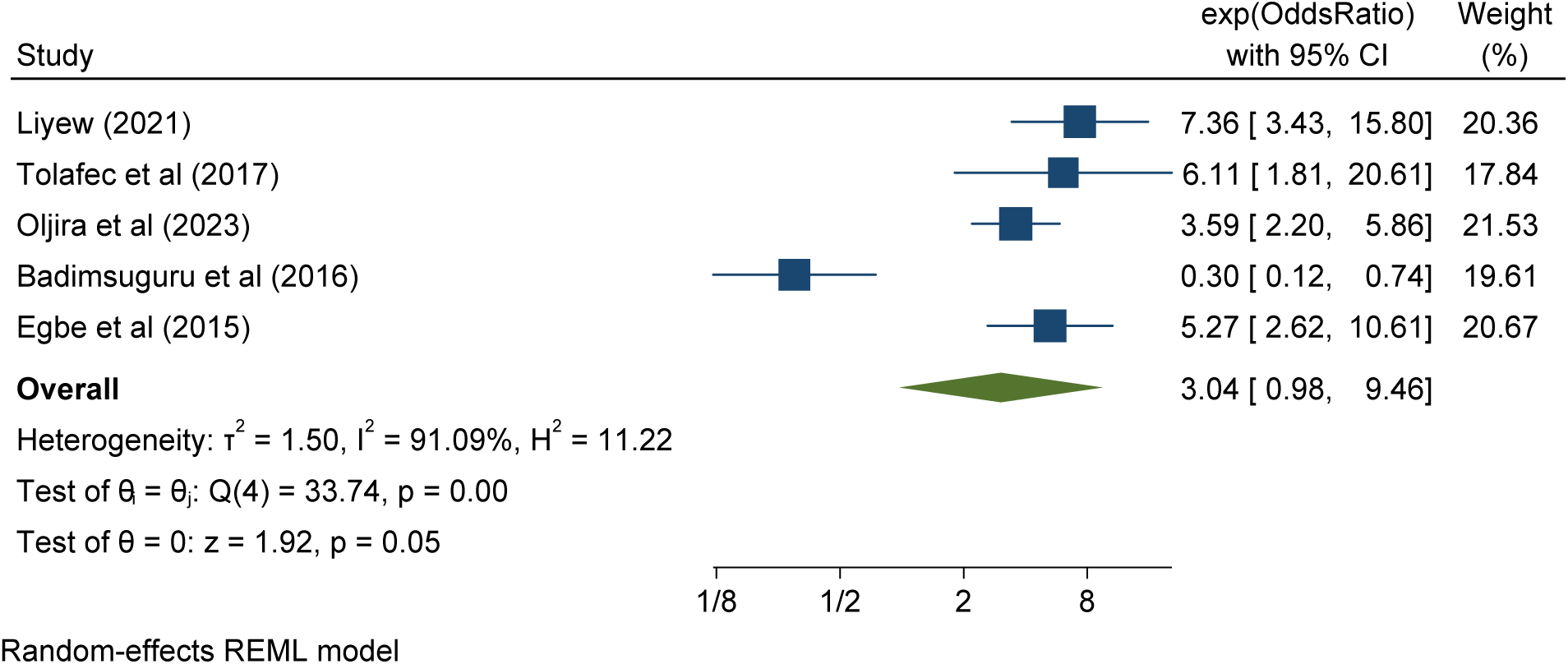
Pooled OR for LBW.

One study from Ethiopia [29] Table 1.0 assessed unique predictors, including maternal syphilis, Rhesus status, and unknown foetal presentation during ANC. As these variables were not evaluated in other included studies, they could not be pooled; however, we have reported the results descriptively.

### Cross sectional study’s findings

A meta-analysis of two studies showed that the parity was significantly associated with the outcome (perinatal mortality), with a pooled odds ratio of 3.96 (95% CI: 2.10–7.47; p < 0.001). Heterogeneity was low (I² = 0.0%), indicating consistent effect sizes across studies.

### Cohort Study’s findings

A random-effects meta-analysis of three studies found that the risk factor was significantly associated with the outcome (pooled RR = 2.30, 95% CI: 1.18–4.52). Study-specific RRs (due to history of stillbirth) ranged from 1.40 (95% CI: 0.72–2.71) in Abdoulaye Hama Diallo Nicolas (22) to 3.37 (95% CI: 1.36–8.34) and 3.60 (95% CI: 1.08–11.94) in Nkwabong, Djientcheu Deugoue (26) for history of IUFD and ENND groups, respectively, with weights between 22.5% and 44.9%. Heterogeneity was moderate (I² = 41.4%, τ² = 0.15), and the Cochran Q-test showed no significant heterogeneity (Q = 3.26, p = 0.20). The overall effect was statistically significant (z = 2.43, p = 0.01).

## Case Control Studies

### Pooled OR for Rural Residence

Two case–control studies from Rwanda [35] and Ethiopia [31] assessed rural residence as a predictor of perinatal mortality. Both studies demonstrated significant associations, with AORs of 3.31 (95% CI: 1.43–7.61) and 2.35 (95% CI: 1.44–3.83), respectively. The pooled random-effects meta-analysis yielded a combined OR of 2.56 (95% CI: 1.68–3.91), indicating that rural residence was associated with more than a twofold increased odd of perinatal death. No evidence of heterogeneity was detected across the studies.

### Pooled OR for Referral

Two case–control studies from Cameroon [24, 25] evaluated the association between referral status and stillbirths. Both studies demonstrated significant associations, with AORs of 3.95 (95% CI: 1.89–6.58) and 14.86 (95% CI: 3.35–66.01), respectively. The pooled random-effects meta-analysis showed a combined OR of 6.90 (95% CI: 2.71–17.52). However, there was evidence of substantial heterogeneity (Q = 2.89, p = 0.089; I² = 65%), suggesting variation in the strength of association across the studies.

### Pooled OR for APH

Two case-control studies [24, 30] assessed the association of APH and placental abruption with stillbirths. Both studies demonstrated strong associations, with AOR of 5.74 (95% CI: 2.67– 12.33) and 21.46 (95% CI: 2.36–194), respectively. A random-effects meta-analysis yielded a pooled OR of 7.28 (95% CI: 2.70–19.65), indicating that women with these complications had more than sevenfold increased odds of stillbirth compared to those without. Heterogeneity across the studies was low (Q = 1.23, I² ≈ 0%), suggesting consistent findings across settings.

### Pooled OR for Congenital Anomalies

Two case–control studies [25, 31] assessed congenital malformations as a predictor of perinatal mortality. The studies reported adjusted odds ratios of 9.15 (95% CI: 2.98–28.0) and 13.0 (95% CI: 1.10–165.3), respectively. A random-effects meta-analysis yielded a pooled OR of 9.7 (95% CI: 3.5–27.0), indicating nearly a tenfold increase in odds of perinatal death among infants with congenital malformations. There was no evidence of heterogeneity (Q = 0.06, I² = 0%).

### Pooled OR for Induced labour

Two case–control studies from Ethiopia [30, 31] assessed the association between induced labour and perinatal mortality. Both studies showed increased odds of adverse outcomes, with AORs of 2.12 (95% CI: 1.09–4.11) and 2.23 (95% CI: 1.33–3.72), respectively. The pooled random-effects analysis yielded a combined OR of 2.19 (95% CI: 1.46–3.29), indicating that induction of labour was associated with more than a twofold increase in the odds of perinatal death or stillbirth. There was no evidence of heterogeneity (Q = 0.01, I² = 0%).

### Pooled OR for Dystocia

Two case–control studies from Ghana [36] and Cameroon [24] assessed the association of labour complications with stillbirths. Prolonged labour was associated with an AOR of 3.5 (95% CI: 1.94– 6.61), while dystocia had an AOR of 5.31 (95% CI: 1.77–15.92). A pooled random-effects meta-analysis yielded a combined OR of 3.86 (95% CI: 2.26–6.60), indicating that labour complications were associated with nearly a fourfold increase in the odds of stillbirth. There was no evidence of heterogeneity (Q = 0.42, I² = 0%).

### Pooled OR for non-use of Partograph Use

Two case–control studies from Ghana [36] and Cameroon [24] examined labour complications and the risk of stillbirth. Prolonged labour was associated with an AOR of 3.50 (95% CI: 1.94– 6.61), while dystocia had an AOR of 5.31 (95% CI: 1.77–15.92). The pooled random-effects analysis showed an overall OR of 3.86 (95% CI: 2.26–6.60), indicating that labour complications increased the odds of stillbirth nearly fourfold. No heterogeneity was observed (Q = 0.42, I² = 0%).

### Pooled OR for Prematurity

Two case–control studies from Ethiopia [31] and Cameroon [24] examined prematurity as a predictor of perinatal mortality. The studies reported AORs of 2.75 (95% CI: 1.61–4.70) and 2.81 (95% CI: 1.48–5.35), respectively. The pooled random-effects analysis yielded a combined OR of 2.77 (95% CI: 1.81–4.26), indicating that prematurity was associated with nearly a threefold increased odd of perinatal death or stillbirth. No heterogeneity was observed (Q ≈ 0.01, I² = 0%).

### Pooled OR for LBW

Five case–control studies conducted in Ethiopia, Cameroon, and Ghana assessed the association between LBW and perinatal mortality or stillbirths. Individual study estimates varied, with AORs ranging from 0.30 (95% CI: 0.12–0.74) in the study by Badimsuguru, Nyarko (36) to 7.36 (95% CI: 3.43–15.81) in the study by Liyew, Molla (30). Other studies also reported elevated risks, including Tolefac, Halle-Ekane (37) with an AOR of 6.11 (95% CI: 1.81–20.59), Oljira, Merdassa (31) with an AOR of 3.59 (95% CI: 2.20–5.87), and Egbe, Ewane (24) with an AOR of 5.27 (95% CI: 2.62–10.62). A random-effects meta-analysis produced a pooled OR of 3.04 (95% CI: 0.98– 9.46), suggesting that infants with LBW had nearly a threefold increased odd of perinatal death or stillbirth compared to normal birth weight infants.

### Certainty of Evidence

Based on the GRADE approach, the overall certainty of evidence for the pooled estimates of perinatal mortality in sub-Saharan Africa was judged to be low to moderate. This rating reflects concerns regarding study quality (risk of bias in observational designs), statistical heterogeneity, and imprecision of some estimates. Nevertheless, the consistency of associations across multiple studies strengthens confidence that the observed findings represent a true underlying burden.

## Discussion

The major predictors of perinatal mortality identified in this review, based on pooled effect estimates, were parity in the two cross-sectional studies and history of stillbirth, intrauterine foetal death (IUFD), and ENND in the three cohort studies. Case–control studies formed the largest group, with pooled effect measures available for two studies each on rural residence, referral status, APH, foetal congenital anomalies, induced labour, dystocia, non-use of the partograph, and prematurity. LBW was the most frequently reported predictor, with five case–control studies contributing to the pooled analysis. Geographically, most studies were conducted in East Africa (Ethiopia, Rwanda, Sudan, and Uganda), followed by Central Africa (three studies from Cameroon), and one study each from Burkina Faso and Ghana. Outcomes also varied, with seven studies reporting stillbirth, six reporting perinatal death, and only one focusing specifically on early neonatal deaths. Although one study by Agena and Modiba [29] could not be included in the pooled analyses, its findings highlight additional risk factors—such as maternal syphilis, Rhesus status, and foetal presentation during antenatal care—that remain underexplored in the wider literature. These results underscore the need for future research to systematically assess such predictors across diverse populations in SSA.

The association between a history of perinatal mortality and subsequent risk of perinatal death observed in this study is consistent with previous evidence, which has consistently identified prior perinatal loss as a strong predictor compared with mothers without such a history [14, 38–40]. Similarly, our findings on APH, foetal congenital anomalies, dystocia, and induced labour align with earlier studies reporting that maternal medical conditions and pregnancy or intrapartum complications—including APH, congenital anomalies, and obstructed labour—are strongly associated with perinatal mortality [41–43]. In terms of intrapartum monitoring, this review highlighted non-use of the partograph as a significant predictor of perinatal mortality. This finding is strongly corroborated by other studies linking failure to use labour monitoring tools with higher risks of perinatal death [44–46]. Prematurity also emerged as a consistent high-risk factor, while the most striking predictor was LBW. This is in line with prior literature demonstrating that both prematurity and LBW are independent and powerful predictors of perinatal mortality compared with term deliveries and normal birthweight infants [14, 39, 47, 48].

In this review, rural residence was found to be significantly associated with perinatal mortality. Interestingly, Lisonkova, Haslam (49) reported that while rural residence was strongly linked to neonatal morbidity, perinatal mortality did not differ between rural and urban groups. Referral status also demonstrated a pooled effect on perinatal mortality, consistent with earlier studies identifying referral as an important contributory factor. Inefficient and poorly structured referral systems in low- and middle-income countries (LMICs) have frequently been cited as critical contributors to perinatal deaths [46, 50–52]. Beyond the pooled effect estimates, this review also identified a number of additional risk factors for perinatal mortality in sub-Saharan Africa, reported across individual studies. For clarity, these predictors can be grouped into four domains.

The sociodemographic determinants included advanced maternal age, low maternal education (secondary level or below), poor household socioeconomic status, lack of access to radio as a source of health information, and seasonal variation, with some studies showing higher risks during the dry season. These findings are consistent with previous evidence indicating that women of low socioeconomic status, those with little or no education, and those residing in rural areas are more vulnerable to perinatal mortality than their wealthier, educated, or urban counterparts [42, 46, 53]. More recently, a 2024 analysis of Demographic and Health Survey (DHS) data from 27 SSA countries found that rural residence and lower household wealth were significantly associated with increased perinatal mortality, while higher maternal education and greater household income were protective against perinatal death [54].

Maternal complications and behavioural factors identified in this review included high gravidity (>5), anaemia during pregnancy, preeclampsia, alcohol consumption, both short and long interpregnancy intervals, and severe maternal outcomes. Biological determinants, such as Rhesus incompatibility and syphilis infection, also emerged as important maternal-level risks. These findings are consistent with evidence from previous studies in similar settings. For example, grand multiparity has been shown to increase the risk of perinatal mortality [54], while anaemia during pregnancy is associated with nearly a threefold higher risk of low birthweight, prematurity, and perinatal death [55]. Preeclampsia and other hypertensive disorders are also well-established contributors, with studies from India and Tanzania confirming particularly high risks [56]. Similarly, prenatal alcohol exposure has been associated with an increased risk of miscarriage, stillbirth, and low birth weight, with broader evidence linking maternal alcohol consumption to a wide range of adverse perinatal outcomes. [57–59]

With respect to interpregnancy interval, our review found both short and long intervals were associated with elevated risk of perinatal mortality. This contrasts with more recent large-scale analyses, which primarily reaffirm short intervals (<18–24 months) as a significant risk and support the WHO birth-spacing guidance [60, 61]. Severe maternal outcomes also emerged as strong predictors in our study, consistent with prospective data from sub-Saharan Africa showing that maternal near-miss events are associated with a fourfold increased risk of adverse perinatal outcomes, including stillbirth and neonatal death [61, 62]. Evidence on Rhesus incompatibility likewise confirms substantial perinatal mortality and foetal death risks due to haemolytic disease of the foetus and newborn, even in the presence of treatment [63]. Finally, our findings on maternal syphilis are in line with meta-analyses and WHO estimates that demonstrate strong associations with stillbirth, neonatal death, and other adverse outcomes. Recent surveillance studies further show persistently high stillbirth rates in pregnancies complicated by congenital syphilis [64, 65].

Our findings mirror a broad body of evidence from SSA and other LMICs. Home delivery, usually with hardly any continuous intrapartum monitoring, is consistently associated with higher perinatal mortality than facility birth [66]. Caesarean delivery, while lifesaving when promptly and appropriately done, is linked to markedly poor perinatal outcomes in many LMIC settings when performed emergently or late in labour, reflecting underlying complications and delays in care; pooled analyses report multiple-fold higher perinatal mortality for emergency compared to elective (scheduled) operations [67, 68]. This study also found breech presentation, abnormal lie, and unknown foetal presentation to be associated with perinatal mortality. Risk from malpresentation are well documented, with recent analyses highlighting elevated mortality in births complicated by breech and transverse/oblique lie [69].

Delayed or inadequate antenatal care—evidenced by late ANC booking, fewer than three ANC contacts were other findings noted in this review. The association between delated or inadequate ANC and poor perinatal outcomes including perinatal deaths is consistent WHO guidance advocating at least eight ANC contacts to reduce stillbirths and adverse outcomes, and with recent studies linking late booking or few contacts to worse maternal-newborn results in SSA [70, 71]. A prior history stillbirth or perinatal was also consistently associated with poor outcomes including perinatal death. Other authors likewise reported that history of poor perinatal outcomes predicts recurrence, as shown in meta-analyses of subsequent pregnancies [72, 73]. On foetal and neonatal predictors, the dominance of LBW and prematurity align with global syntheses signalling that majority of neonatal deaths occur among LBW infants, over half of whom are preterm [4]. Additional intrapartum and ENND predictors, meconium-stained liquor, birth asphyxia, and RDS, are repeatedly linked to death in settings with limited resuscitation and intensive care capacity [74, 75].

Our review also highlighted neonatal infections as a predictor of perinatal mortality in one study. This finding aligns with global evidence identifying sepsis as a leading direct cause of neonatal deaths, responsible for roughly over 15% of neonatal mortality worldwide, with the highest burden in SSA and South Asia [76, 77]. Evidence from systematic reviews shows that simple, low-cost measures, such as clean delivery practices, cord care, early breastfeeding, and timely antibiotics, can substantially reduce infection-related deaths [78]. Lastly, failure to initiate breastfeeding promptly and the intrinsic risk of multiple gestation, most notable in twins, are well supported by meta-analyses and regional data, with delayed breastfeeding initiation associated with substantially higher mortality and twins experiencing several-fold higher infant mortality than singletons [79, 80].

### Study implications

Based on pooled estimates, key sociodemographic predictors included rural residence and referral status. Additional predictors, though not pooled, were advanced maternal age, low education level, poor household socioeconomic status, lack of access to communication channels such as radio, and seasonal variation (with higher risks during the dry season). Maternal and obstetric complications associated with perinatal mortality included APH, foetal congenital anomalies, previous history of perinatal death, induced labour, dystocia, non-use of the partograph, prematurity, and LBW. Other predictors identified, though not meta-analysed, were maternal syphilis, Rhesus incompatibility, foetal malpresentation, and delayed or inadequate antenatal care.

These findings highlight the need for multifaceted and targeted policy interventions that address the identified sociodemographic and economic disparities. Strengthening context-specific information, education, and communication (IEC) strategies is essential to influence health-seeking behaviours. In parallel, maternal, obstetric, and neonatal care must be improved across the continuum of care—including prenatal, antenatal, intrapartum, and newborn services. Beyond contributing to the body of knowledge, this review underscores the urgent need for further research and the development of targeted implementation strategies to advance maternal and newborn health outcomes in the region.

### Strengths and limitations

This systematic review and meta-analysis provide a comprehensive synthesis of evidence on perinatal mortality in sub-Saharan Africa over the past two decades. By pooling effect estimates from diverse observational studies, it highlights the persistent burden of perinatal deaths in the region and identifies key predictors across sociodemographic, maternal, obstetric, and neonatal domains. The findings not only underscore the multifactorial nature of perinatal mortality but also offer valuable insights to guide targeted interventions and inform policies aimed at improving maternal and newborn health. Importantly, the thematic organisation of predictors provides a structured framework for comparison with evidence from other regions and contributes to a deeper understanding of context-specific drivers of perinatal mortality.

Nevertheless, some limitations must be acknowledged. Subgroup analyses were planned to explore sources of heterogeneity, but their interpretation was constrained by the small number of studies in certain regions, particularly West Africa (two studies) and Central Africa (three studies). As most included studies (nine of 14) were from East Africa, important regional differences and context-specific predictors may have been underrepresented. Furthermore, variability in study designs, outcome definitions, and measurement of exposures may have introduced heterogeneity, limiting the precision of pooled estimates. Future research should therefore aim for a more balanced geographical representation and employ harmonised definitions and methodologies to enable more robust subgroup analyses and stronger regional comparisons.

### Conclusion

The principal predictors of perinatal mortality identified through pooled estimates included parity, history of perinatal mortality, rural residence, referral status, APH, foetal congenital anomalies, induced labour, dystocia, non-use of the partograph, and prematurity. LBW emerged as the most frequently reported risk factor, with five studies contributing to the pooled estimate. Several additional determinants could not be synthesised quantitatively but were consistently reported, including sociodemographic factors such as low maternal education and poor household income, as well as maternal factors such as anaemia, preeclampsia, Rhesus incompatibility, interpregnancy interval, and infections.

These findings underscore the multifactorial nature of perinatal mortality in sub-Saharan Africa and highlight the complex interplay of social, maternal, obstetric, and neonatal risks. Addressing this burden requires integrated strategies that strengthen antenatal, intrapartum, and neonatal care while tackling the broader social determinants that place mothers and infants at risk.

### Recommendations

This study underscores the urgent need to strengthen referral systems and expand access to high-quality maternal and neonatal health services, particularly at facilities equipped with Comprehensive Emergency Obstetric and Neonatal Care (CEmONC). Such facilities are uniquely positioned to provide both advanced obstetric interventions and essential neonatal care. Efforts should prioritize early ANC booking, alongside the delivery of adequate, evidence-based ANC that facilitates timely detection and management of high-risk maternal, obstetric, and foetal conditions. These clinical improvements must be complemented by targeted interventions aimed at reducing or preventing the identified predictors of perinatal mortality.

Beyond facility-based interventions, the contribution of behavioural, socioeconomic, and demographic determinants highlights the need for robust community-based strategies. Tailored IEC initiatives, coupled with empowerment programs, are essential to equip girls and women with the knowledge, skills, and resources required to ensure safe pregnancies and positive birth outcomes. Addressing these structural and behavioural barriers will be critical to reducing preventable perinatal deaths and advancing progress in maternal and newborn health across sub-Saharan Africa.

## Data Availability

This study is based on secondary data derived from publicly available published sources. All data used in the analysis are cited within the manuscript and can be accessed through the original publications. No new data were generated or collected specifically for this study.

## Acknowledgements

I would like to thank all my supervisors for the guidance provided to complete this study. I also would like to acknowledge the immense contribution made by the research assistants that handled and managed data.

## Authors’ contributions

MM: Study conceptualisation, literature review, data management, meta-analysis, and manuscript writing. PK supervision and methodology. CJ: supervision and study contextualisation KS: data search MWM: data search. BV: Study conceptualisation, contextualisation, and overall supervision. All authors read and approved the final version of the manuscript.

